# Age-related white matter differences in children with neurofibromatosis type 1 compared to controls

**DOI:** 10.1101/2023.09.29.23295837

**Authors:** L. Bruckert, K. E. Travis, L. T. Tam, K. W. Yeom, C. J. Campen

## Abstract

Neurofibromatosis type 1 (NF1) is a common genetic condition in which 30-70% of children experience learning challenges including deficits in attention, executive function, and working memory. White matter pathways have been implicated in these cognitive functions; yet, they have not been well characterized in NF1. In this retrospective cohort study, we used diffusion MRI tractography to examine the microstructural properties of major white matter pathways in 20 children with NF1 aged 1 year to 18 years relative to 20 age-and sex-matched controls. An automated approach was used to identify and extract mean diffusivity (MD) and fractional anisotropy (FA) of eight cerebral white matter pathways bilaterally and the anterior and posterior part of the corpus callosum. Compared to controls, children with NF1 had significantly increased MD and significantly decreased FA in multiple white matter pathways including the anterior thalamic radiation, cingulate, uncinate fasciculus, inferior fronto-occipital fasciculus, arcuate fasciculus, and corticospinal tract. Differences in MD and FA remained significant after controlling for intracranial volume. In addition, MD and FA differences between children with NF1 and controls were greater at younger than older ages. These findings have implications for understanding the etiology of the neurocognitive deficits seen in many children with NF1.

## 1 Introduction

Neurofibromatosis type 1 (NF1) is one of the most common genetic disorders affecting approximately 1 in 3000 children.^1^ NF1 prevalence is not affected by sex, race, or ethnicity. Clinical manifestations vary greatly in presentation and severity and affect nearly every organ system in the body. They include changes in pigmentation like café-au-lait macules, skinfold freckling, or Lisch nodules, vasculopathy, including renal artery stenosis or moyamoya syndrome, macrocephaly, and nervous system tumors like neurofibromas or gliomas and other cancers. Children with NF1 also exhibit high rates of behavioral and learning problems that significantly impact their quality of life.^2^ Specifically, 30-70% of children with NF1 experience deficits in executive function, working memory, language, and intellectual abilities more broadly.^2,3^

NF1 is caused by the mutation of a gene on chromosome 17, which hinders the production of the protein neurofibromin, a tumor suppressor that acts on the rat sarcoma (Ras) pathway.^4^ Loss of neurofibromin and subsequent increase in Ras signaling has been shown to affect neuronal cell differentiation, growth, and apoptosis relevant to normal brain development.^5–7^ In line with these pathophysiological changes in neurons’ cellular processes, neuroimaging studies have revealed various brain abnormalities in NF1. These include lower cortical gyrification,^8^ focal hyperintense lesions on T2-weighted images, the so-called “unidentified bright objects”, ^9,10^ and widespread volumetric abnormalities.^11^ For example, grey and white matter volume increases have been consistently documented in children and adolescents with NF1 relative to healthy peers.^11–13^ Similarly, the volume and thickness of the corpus callosum is increased in NF1.^13,14^ The origins and significance of these brain abnormalities, however, remain unclear.

Other frequently observed abnormalities are diffuse reductions of white matter integrity.^15–17^ Diffusion magnetic resonance imaging (MRI) is commonly used to assess white matter properties *in vivo.*^18^ By measuring the movement of water molecules throughout the brain, diffusion MRI can provide unique insights into tissue microstructure that are otherwise not detected using conventional MRI.^19,20^ The most common metrics derived from diffusion MRI are mean diffusivity (MD) and fractional anisotropy (FA). MD indexes the overall magnitude of water diffusion; FA indexes the restriction of water diffusion in a particular direction. Diffusion MRI, analyzed using tractography, allows for the interrogation of white matter properties of specific white matter pathways.^18,21^ Several tractography methods have been developed to perform the reconstruction of white matter pathways within native space, thus providing increased anatomical precision compared to ROI or whole-brain analysis methods.

To date, only a few studies have examined diffusion MRI metrics in NF1. Most of these studies used a region-of interest approach and found significantly increased MD in both hyperintense lesions and in the normal-appearing brain areas in children and adults with NF1 compared to healthy peers.^22–25^ Other studies used a whole-brain approach and found widespread white matter alterations as indexed by significantly increased MD and significantly decreased FA in adolescents and young adults with NF1.^16,26,27^ Only one group has used diffusion MRI tractography to reconstruct and investigate a specific white matter pathway, namely the optic pathway in NF1.^26,28^ In their studies, de Blank and colleagues demonstrated that i) a decrease in FA of the optic radiations was associated with visual acuity loss in children with NF1 with optic pathway gliomas^28^ and ii) that age-related change of diffusion MRI metrics of the optic radiations was different in children with NF1 relative to peers.^26^ The latter was assessed using multiple regression analysis that examined the effect of ln(age), sex, NF1 status, and the interaction term ln(age) by NF1 status on diffusion MRI metrics. The interaction term ln(age) by NF1 status reached statistical significance in this cross-sectional study indicating that diffusion metrics of the optic radiations were reduced in young children with NF1 and matured more slowly compared to children without NF1.^26^ A similar effect of age on diffusion MRI metrics in children with NF1 was shown by Tam and colleagues.^24^ They found that FA was reduced in the corpus callosum and frontal white matter areas in children with NF1 relative to healthy peers.^24^ Strikingly, the differences in FA were predominantly driven by the early and middle childhood age groups but not the adolescent one. These findings suggest potential differences in the developmental trajectory of the optic radiations and other white matter areas due to NF1.

While white matter alterations were observed throughout the brain, to our knowledge no one has reported diffusion MRI metrics controlling for known differences in intracranial volume in children with NF1 relative to controls. In addition, no study has used diffusion MRI tractography to systematically evaluate microstructural properties of white matter pathways across children, adolescents, and young adults with NF1. Compared to conventional region of interest methods, diffusion MRI tractography allows the identification of the entire white matter pathway. It is therefore less susceptible to interrogator bias and volume averaging.

In this study, we aim to describe a comprehensive set of major white matter pathways in children with NF1 across a broad age range. Typical development of specific white matter pathways is critical for normal development of cognitive functions. Thus, our findings are important for understanding how NF1 may impact white matter development and how alterations in white matter development may contribute to cognitive outcomes in NF1. We use an automated tractography method, namely automated fiber quantification (AFQ),^29^ to segment and characterize nine white matter pathways in children with NF1 aged 1 year to 18 years compared to age-and sex-matched controls. AFQ allows us to identify white matter pathways in children’s native space and has been shown to reliably segment white matter pathways across a wide age-range of children.^30,31^ Based on previous findings, we hypothesize that i) children with NF1 would have higher MD and lower FA across many major white matter pathways^16,27^ and that ii) the magnitude of these differences would be affected by the age, such that MD/FA differences between children with NF1 and controls would be more pronounced in younger than older children.^24,26^

## 2 Methods

### 2.1 Participants

A database of pediatrics patients, who were treated at Lucile Packard Children’s Hospital at Stanford between January 2010 and November 2017, was queried retrospectively after institutional review board approval (protocol 28674). *Figure 1* depicts the consort flow diagram for inclusion and exclusion of pediatric patients. Twenty-nine children met the following inclusion criteria: i) confirmed NF1 diagnosis, ii) no medical history of moyamoya; iii) no medical history of optic pathway glioma; iv) no other intracranial mass; v) no systemic chemotherapy. We excluded six children that were not scanned at 3T using high resolution T1- weighted volumetric and diffusion MRI. We excluded three more patients whose diffusion scans were affected by technical issues including poor whole-brain coverage (n = 2) and large signal dropouts (n = 1). Our final sample included 20 children with NF1 and 20 age-and sex-matched healthy controls (*Table 1*). Healthy controls were taken from the same database and included children who were scanned at the hospital for a clinical indication (e.g., isolated headaches, nausea, scalp nevus, peri-orbital dermatoid, facial hemangioma, benign strabismus without orbital or intracranial abnormality, sinus disease or inflammatory nasal obstruction, ear infection, syncope without a history of generalized seizures, and family history of aneurysm or vascular malformations) but whose brain scans were read as normal and follow-up medical assessments did not lead to a clinical diagnosis. The control cohort has been described in detail by Bruckert and colleagues.^30^

**Figure 1.**
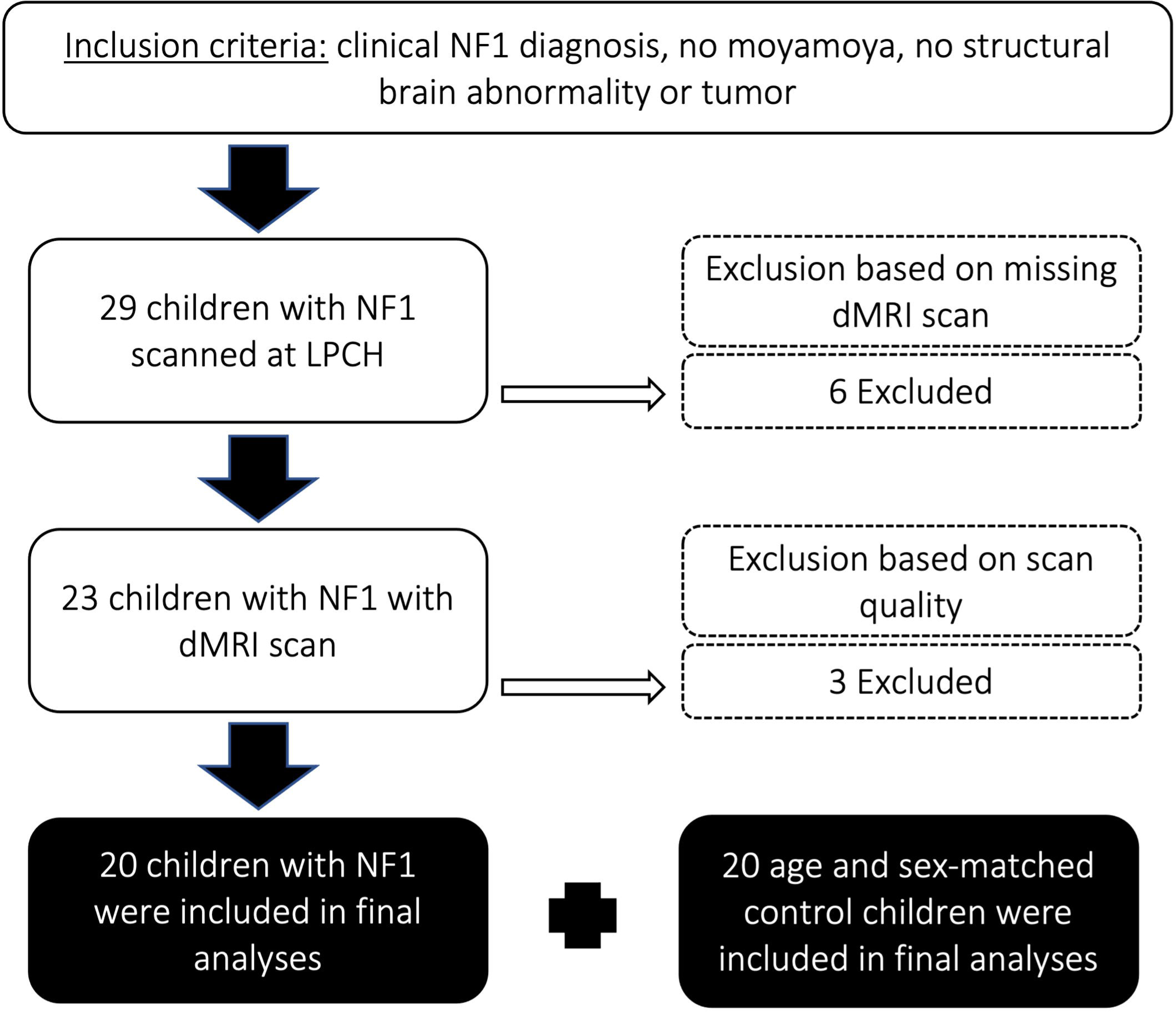
Consort flow diagram describing inclusion and exclusion criteria of pediatric patients in this study. NF1 = Neurofibromatosis Type 1; LPCH = Lucile Packard Children’s Hospital Stanford; dMRI = diffusion magnetic resonance imaging

**Table 1.**
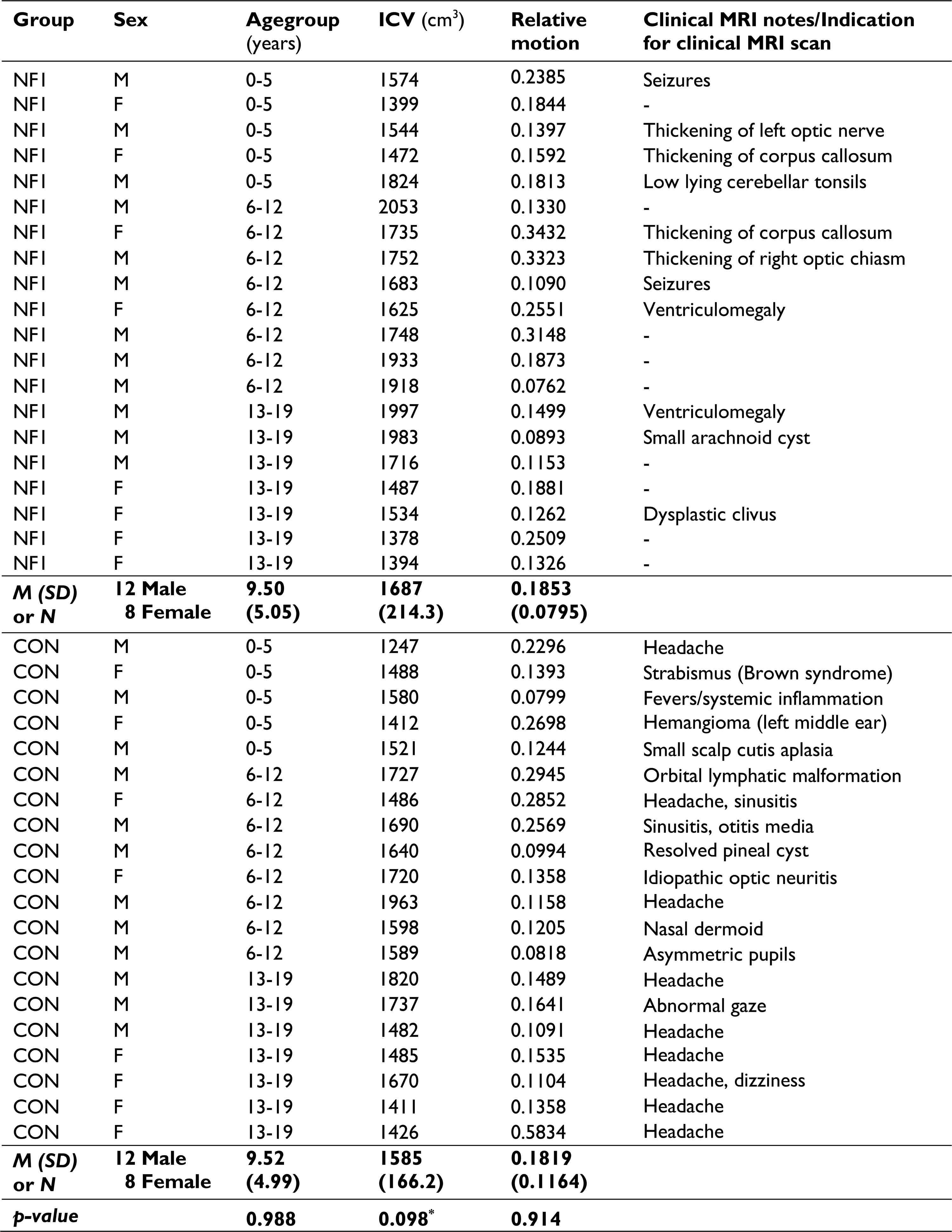
Characteristics of the sample (p-values < 0.1 are marked by an asterisk).

### 2.2 MRI data acquisition and preprocessing

Imaging parameters and methods for dMRI preprocessing and analyses of relative head motion have been described in previous publications^30,32^ and are briefly summarized below.

MRI data were obtained at 3T (GE MR750 Discovery; GE Healthcare, Waukesha, WI, USA) with an 8-channel head coil. Children between 3 months and 6 years of age were sedated under general anesthesia, and some children aged 6 years and older were sedated based on individual maturity level and ability to tolerate the MRI exam. Both high-resolution T1- weighted (3D SPGR, TR= 7.76 ms, TE = 3.47 ms, FOV= 240 × 240 mm2, acquisition matrix = 512 × 512, voxel size = 0.4688 × 0.4688 × 1 mm3, orientation = axial) and diffusion-weighted images were acquired as part of the pediatric brain MRI protocol.

High-resolution T1-weighted images were used for aligning low resolution diffusion MRI scans and to calculate the intracranial volume (ICV) for each child. FMRIB’s Automated Segmentation Tool (FAST)^33^ was used to segment the T1-weighted images into three different tissue types, namely grey matter, white matter, and cerebrospinal fluid whilst also correcting for spatial intensity variations. The resulting maps of grey matter volume, white matter volume and cerebrospinal fluid volume were used to calculate ICV.

Diffusion data were collected with a twice-refocused GRAPPA DT-EPI sequence (TR= 4000–6000 ms depending on slice coverage, TE = 76.59 ms, FOV= 240 × 240 mm^2^, acquisition matrix = 256 × 256, voxel size = 0.9375 × 0.9375 × 3 mm^3^) using a b-value of 1000 s/mm^2^ sampling along 25 isotropically distributed diffusion directions. One additional volume was acquired at b = 0 at the beginning of each scan.

We used the open-source software mrDiffusion (https://github.com/vistalab/vistasoft/tree/master/mrDiffusion) implemented in MATLAB R2014a (Mathworks, Natick, MA, United States) to preprocess the diffusion data. The non-diffusion image (b0) was registered to the subject’s T1-weighted image, which had been aligned to the canonical ac-pc orientation. Diffusion-weighted images were corrected for head motion by rigid-body registration to the b0 image.^34^ Relative head motion was assessed for each subject following the procedure described by Bruckert and colleagues.^30^ No subjects were excluded due to excessive motion. The combined transform that resulted from the alignment to the T1-weighted image and motion correction was applied to the diffusion data once, and the transformed images were resampled to 2 × 2 ×2 mm^3^ isotropic voxels. Diffusion gradient directions were adjusted to fit the resampled diffusion data.^35^ Using a standard least-squares algorithm, maps of MD and FA were generated.

### 2.3 White matter tract identification

The open-source software, Automated Fiber Quantification (AFQ),^29^ was used to track and segment cerebral white matter pathways in each child’s native space. Tractography was seeded from each voxel in a white matter mask (FA > 0.2) and deterministic tracking proceeded in all directions until FA values dropped below 0.15, or until the angle between the last path segment and next step direction was greater than 30°. Segmentation of the bilateral anterior thalamic radiation (ATR), corticospinal tract (CST), cingulate (Cing), inferior fronto-occipital fasciculus (IFOF), inferior longitudinal fasciculus (ILF), superior longitudinal fasciculus (SLF), uncinate fasciculus (UF), arcuate fasciculus (AF) and the anterior (forceps minor, FMinor) and posterior (forceps major, FMajor) part of the corpus callosum was based on an automated waypoint ROI method implemented in AFQ (see Yeatman et al.^29^ for details). The core of the tract was calculated by defining 30 sample points along the tract and computing the robust mean position of the corresponding sample points. The robust mean was computed by estimating the three-dimensional Gaussian covariance of the sample points and removing fibers that were either more than 5 standard deviations away from the mean position of the tract or that differed more than 4 standard deviations in length from the mean length of the tract. For the SLF, we used a more rigorous cleaning approach and removed fibers that were either more than 4 standard deviations away from the mean position of the tract or that differed more than 1 standard deviations in length from the mean length of the tract. Fiber renderings for each tract and each child were visually inspected prior to any statistical analyses to ensure that each tract conformed to anatomical norms. Using these methods, we were able to identify these cerebral white matter pathways in the majority of children. A small number of children were excluded from each analysis because a tract could not be segmented or did not conform to anatomical norms; excluded were: four (2 NF1) and six children (3 NF1) from the left and right Cing; seven (2 NF1) and two children (1 NF1) from the left and right SLF; two (0 NF1) and five children (2 NF1) from the left and right AF; two children (1 NF1) from the left ILF; one child (1 NF1) from the left IFOF; one child (0 NF1) from the left UF; and one child (0 NF1) from the FMajor. Diffusion properties (MD, FA) were quantified at 30 equidistant nodes along the central portion of each fiber tract bounded by the same two ROIs used for tract segmentation (*Figure 2*). Mean tract-diffusion indices were calculated by averaging MD or FA values of all 30 nodes.

**Figure 2.**
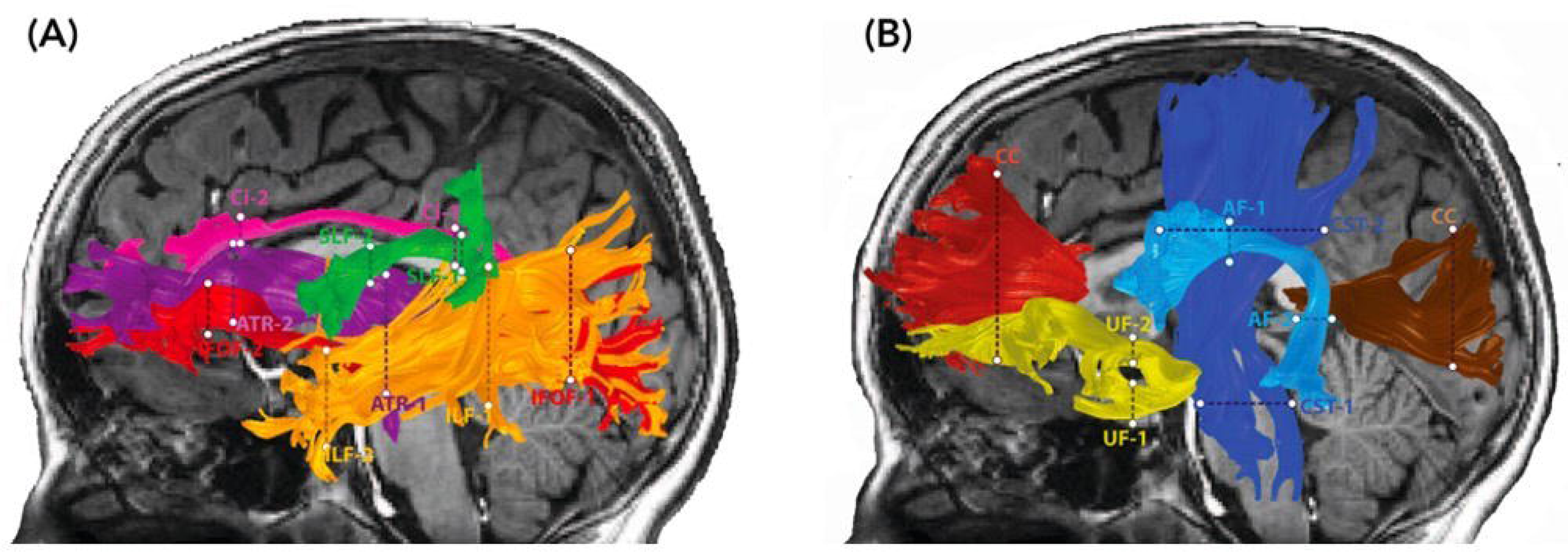
Diffusion MRI tractography of cerebral white matter pathways. Renderings of 10 major white matter pathways are overlaid on two sagittal T1-weighted images of a representative school-aged child with NF1. The defining waypoint regions of interest (ROIs) are marked by dotted lines. The left panel (A) depicts the anterior thalamic radiation (ATR) = purple, superior longitudinal fasciculus (SLF) = green, cingulate (Ci) = pink, inferior fronto-occipital fasciculus (IFOF) = light red, and inferior longitudinal fasciculus (ILF) = orange. The right panel (B) depicts the corticospinal tract (CST) = dark blue, uncinate fasciculus (UF) = yellow, arcuate fasciculus (AF) = light blue, corpus callosum forceps minor (CC) = dark red, and corpus callosum forceps major (CC) = maroon.

### 2.4 Statistical analyses

Statistical analyses were conducted using IBM SPSS software (version 25.0, IBM Corporation, 2014). Statistical significance was set at p < 0.05. Independent t-tests were used to determine whether intracranial volume or relative motion during diffusion MRI differed between children with NF1 and controls. Variables that showed significant group differences were included as covariates in subsequent analyses. To assess the contribution of age to differences in white matter properties (MD and FA) between children with NF1 and controls, we conducted a series of hierarchical multiple regression analyses and included a group (NF1 vs. control) by age (in years) interaction term. If the interaction term reached statistical significance, we performed simple main effetcs analyses to assess the effect of age or group on MD and FA values. We used false discovery rate (FDR, *p* = .05) to account for multiple comparisons of the number of white matter pathways.

### 2.5 Data availability

Qualified investigators may request anonymized data for purposes of replicating procedures and results from the corresponding author.

## 3 Results

Group characteristics of children with NF1 and controls are summarized in Table 1. By design, the groups did not differ in age or distribution of sex. The groups also did not differ in the amount of relative motion during the diffusion MRI scan. Numerically, children with NF1 had larger intracranial volume but the difference did not reach statistical significance. Because previous studies have consistently found that children with NF1 have larger brains than their healthy peers^12,13,16,36^ and because we saw a trend in the same direction in our cohort, intracranial volume was included as a covariate in all regression models to ensure that observed differences in MD and FA were not driven by differences in intracranial volume. Mean tract-MD and FA values for each white matter pathways for children with NF1 and their matched controls are summarized in Table S1.

For mean tract-MD, the regression analyses revealed a significant group-by-age interaction in all white matter pathways, except for the corpus callosum, FMinor, and FMajor (Table 2). Simple main effects analyses showed that children with NF1 had higher MD values compared to age-and sex-matched peers. Differences in MD were also more pronounced at younger than older ages (Figure 3 and Figure S1). For the FMinor, and FMajor, we found a significant main effect of group. Across ages, children with NF1 had higher MD values than controls (Figure 3 and Figure S1).

**Figure 3.**
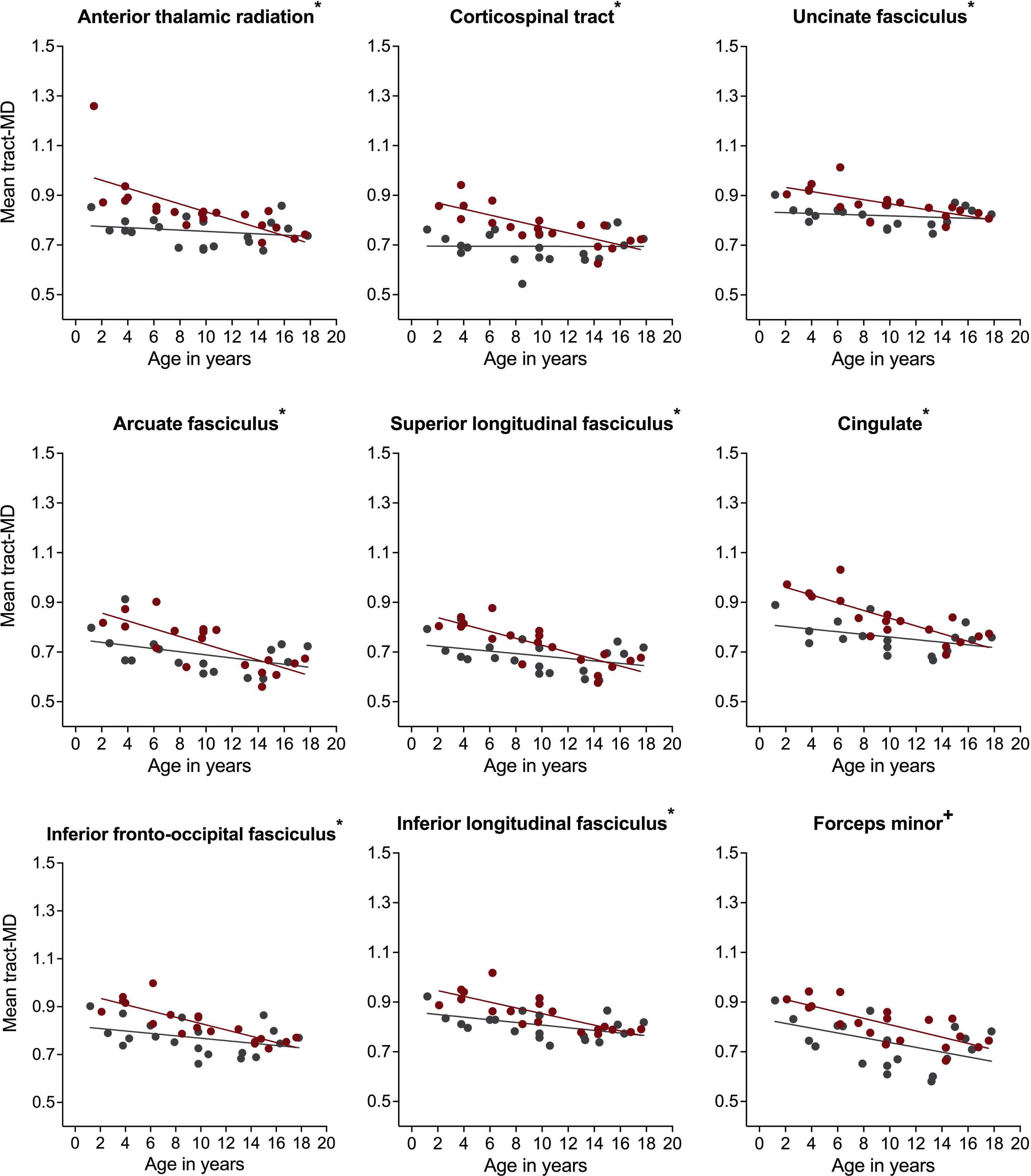
Associations of mean diffusivity (MD) with age in children with neurofibromatosis type 1 (NF1, red circles) compared to age-and sex-matched controls (CON, grey circles) after controlling for intracranial volume. Associations are shown for the <u>right</u> hemisphere only. Graphs marked with an *, +, and ° represent significant group-by-age interaction, significant main effect of group, and significant main effect of age respectively. Results remain significant after correcting for multiple comparisons using false discovery rate of *p* = 0.05.

**Table 2.**
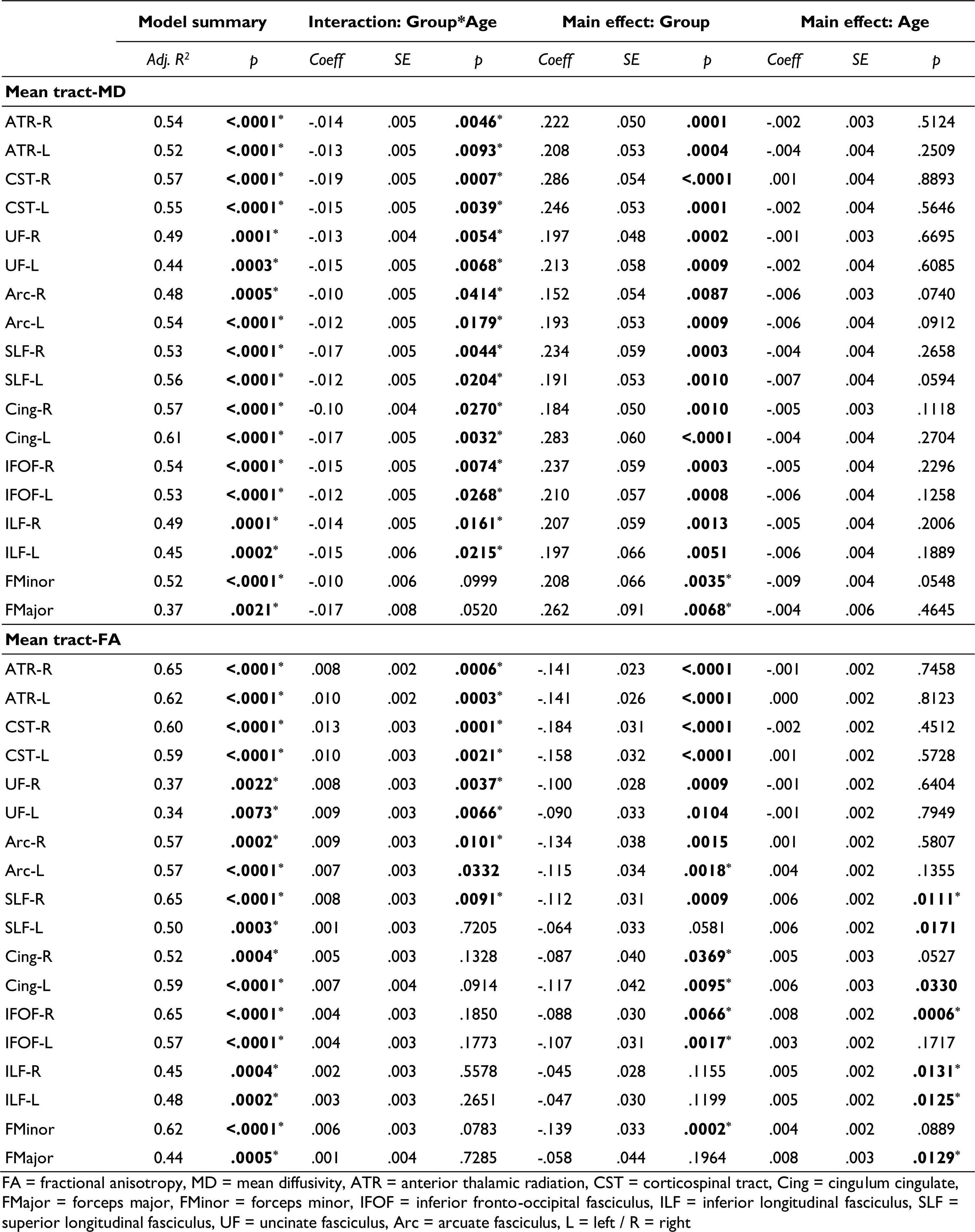
Results from moderation analysis. *P*-values marked with an asterisk remain significant after correcting for multiple comparisons of the number of white matter pathways using a false discovery rate of *p* = 0.05.

For mean tract-FA, analysis revealed a similar pattern of results. We found a significant group-by-age interaction for most white matter pathways including the bilateral ATR, CST, UF, AF, and right SLF (Table 2). Simple main effects analyses showed that children with NF1 had lower FA values compared to controls. Again, these differences were more pronounced at younger than older ages (Figure 4 and Figure S2). For the bilateral Cing, IFOF and FMinor, we found a significant main effect of group with lower FA values in children with NF1 relative to controls across all ages (Figure 4 and Figure S2). For the remaining white matter pathways, namely the bilateral ILF and FMajor, we found a significant main effect of age. Across both groups, FA increased with age. All results remained significant after correcting for the number of white matter pathways using FDR.

**Figure 4.**
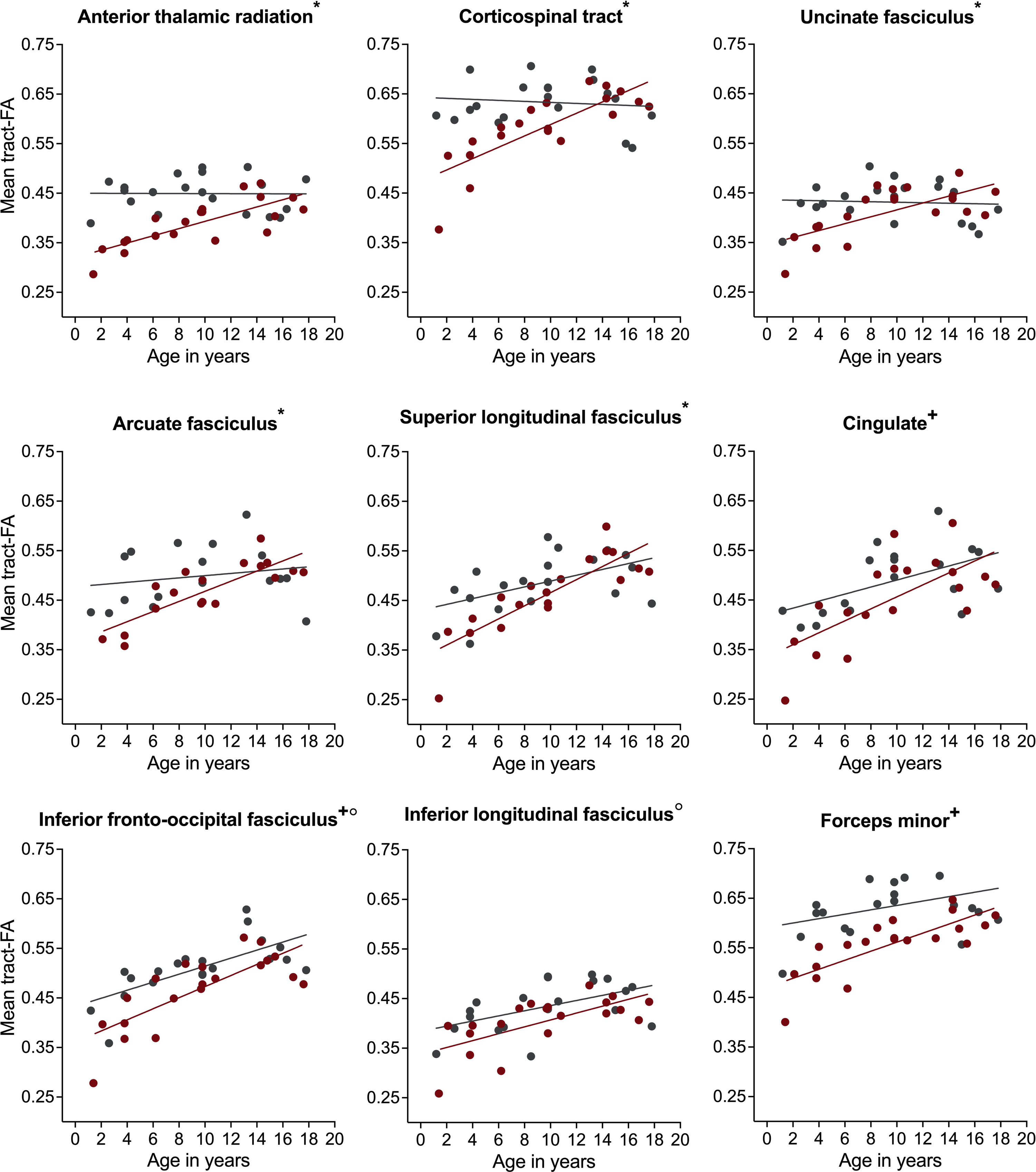
Associations of fractional anisotropy (FA) with age in children with neurofibromatosis type 1 (NF1, red circles) compared to age-and sex-matched controls (CON, grey circles) after controlling for intracranial volume. Associations are shown for the <u>right</u> hemisphere only. Graphs marked with an *, +, and ° represent significant group-by-age interaction, significant main effect of group, and significant main effect of age respectively. Results remain significant after correcting for multiple comparisons using false discovery rate of *p* = 0.05.

## 4 Discussion

Children with NF1 showed differences in microstructural properties of several major white matter pathways compared to age-and sex-matched controls. These differences were widespread and consistently observed in 14 out of 18 white matter pathways as quantified using diffusion MRI tractography. Moreover, these differences persisted when we controlled for intracranial volume. It is important to consider intracranial volume in studies of children with NF1 because individuals with NF1 have larger brains on average.^12,13,16,36^

Our results support previous studies that demonstrated increased MD and decreased FA in children and adults with NF1 compared to controls using whole-brain^16,26^ or ROI analysis ^17,22,23,25^. Karlsgodt and colleagues^16^, for example, found diffuse white matter differences in young adults with NF1 compared to demographically matched controls using a whole-brain analytical approach called tract-based spatial statistics. They also found significantly increased overall grey and white matter volume in the same NF1 patients but did not take these differences into account for their diffusion MRI analysis. To our knowledge this is the first study that demonstrates white matter alterations in a comprehensive set of white matter pathways controlling for overall intracranial volume.

The use of diffusion MRI tractography to examine a white matter pathway in its entirety is another novelty of this study. Only one other study used diffusion MRI tractography in patients with NF1.^28^ In their study, deBlank and colleagues^28^ applied tractography to assess microstructural properties of the optic pathway in children with NF1 with optic pathway gliomas. They found that a decrease in FA of the optic radiations was associated with abnormal visual acuity in children with NF1 with optic pathway gliomas. However, they did not examine other cerebral white matter pathways, assess children without glioma, or include a healthy control group. The use of tractography is advantageous, in that it can be used to reconstruct white matter pathways in native space, without registration to a template. While image registration and normalization methods have been shown to be accurate and reliable in healthy average-sized brains,^37^ they can introduce biases when applied to patient populations with brain abnormalities such as gliomas or varying brain sizes across development.^38,39^ Therefore, we chose tractography as the preferred method to assess white matter properties in our NF1 cohort. Another advantage of tractography is that we obtain a detailed description of the entire pathway of interest. This in turn may aid analyses that focus on brain-function correlations because we can reconstruct and assess white matter pathways that have been previously implicated in specific cognitive functions.

We corroborated findings from our group^24^ and other researchers^26^ that the magnitude of white matter differences between children with NF1 and controls varies by age. Prior reports were limited to ROI analyses of the fronto-temporal white matter, whole-brain analysis, and tractography of the optic pathway only. The current work characterizes a comprehensive set of nine major white matter pathways bilaterally and the corpus callosum throughout the pediatric NF1 brain. Our results indicate that in most white matter pathways, differences were larger at younger ages. While this study did not assess individual children longitudinally, our findings suggest that children with NF1 may have altered or delayed white matter maturation that can lead to persistent white matter abnormalities later in life. Patients with NF1 have previously been shown to have abnormal diffusion metrics.^24,26^ Most diffusion MRI studies, however, have focused on adult patients with NF1^16,17^ ^17,40^ limiting our understanding of NF1-related white matter differences in the developing brain. The definition of normative values for diffusion MRI metrics across ages is an essential step in developing potential diffusion MRI biomarkers, which in turn could facilitate early identification of children with NF1 at risk for neurocognitive or visual deficits. Diffusion MRI studies that have examined children with NF1 have either not assessed the effect of age on diffusion metrics^15^ or focused on whole-brain analysis with a single white matter pathway^16,26^ or were restricted to a set of ROIs.^24^

Results from these and our cross-sectional diffusion MRI study in children with NF1 suggest that white matter differences may not be static throughout a patient’s life but that they change due to maturation and experience-dependent reorganization. Most work on white matter plasticity has focused on myelination, the process of coating axons with a fatty myelin sheath to improve signal conduction.^41,42^ Consistent with postmortem studies,^43^ neuroimaging studies have shown that the most dynamic period of myelination occurs in the first few years of life coinciding with the rapid increase in cognitive abilities.^44,45^ White matter maturation, however, continues well into adolescence and adulthood.^46,47^ Longitudinal studies in children with NF1 are needed to learn if and how NF1-related white matter alterations change over time and to determine if there is a specific window of opportunity during childhood for targeted treatment to delay or moderate any white matter alterations. In addition, future neuroimaging studies should combine diffusion MRI with imaging methods that can provide direct measures of myelin content^48^ to triangulate the underlying biological mechanisms.

The data in this study were analyzed retrospectively and therefore we were limited in our sample size. A sample size of 20 children with NF1 and 20 well-matched controls, however, is comparable (or higher) to previous diffusion MRI studies in NF1.^15,16,27,28^ Further, Lucile Packard Children’s Hospital at Stanford is a tertiary-care pediatric hospital, and our patients may harbor more severe disease than the average child with NF1. However, we excluded children with NF1with moyamoya, glioma, or who had received chemotherapy to limit our sample to children without visible white matter injury or other significant brain abnormalities. In this study, we capitalized on the availability of scans acquired in children who presented at our hospital for a clinical indication as our control cohort. This may make the scans unrepresentative of a community-based sample, however, our thorough chart review (for details see Bruckert et al.^30^) combined with our strict inclusion and exclusion criteria make this possibility unlikely. MRI scans and follow-up reports of all our control participants were normal as reviewed by a pediatric neuroradiologist and developmental-behavioral pediatrician.

## 5 Conclusion

We validated prior findings that white matter microstructure is altered in the pediatric NF1 brain, and that white matter differences are more pronounced in younger children with NF1 compared to healthy peers. Using tractography our study provided increased specificity for a comprehensive set of white matter pathways. In addition, our findings suggest that diffusion MRI abnormalities are due to differences in white matter microstructure rather than differences in brain volume seen in NF1. These differences may point to aberrant oligodendroglial precursor cell dynamics during childhood and may have implication for cognitive development and gliomagenesis in NF1. Children with NF1 harbor high rates of learning challenges including attention, executive function, and processing disorders and high rates of low-grade gliomas. Future studies should investigate the relationship of white matter pathways implicated in cognition or affected by gliomas and neurocognitive and clinical outcome to understand the clinical relevance of the observed white matter alterations.

## Data Availability

All data produced in the present study are available upon reasonable request to the authors.

## 6 Acknowledgements

We would also like to thank E.S. McKenna and K. Shpanskaya for their help curating the MRI cohort data.

## 7 Funding

This work was made possible through a generous donation from the Greathouse Family Foundation.

## 8 Competing interests

The authors report no competing interests.

---

***Figure S1.* Associations of mean diffusivity (MD) with age in children with neurofibromatosis type 1 (NF1, red circles) compared to age-and sex-matched controls (CON, grey circles) after controlling for intracranial volume.** Associations are shown for the <u>left</u> hemisphere only. Graphs marked with an *, +, and ° represent significant group-by-age interaction, significant main effect of group, and significant main effect of age respectively. Results remain significant after correcting for multiple comparisons using false discovery rate of *p* = 0.05.

***Figure S2.* Associations of fractional anisotropy (FA) with age in children with neurofibromatosis type 1 (NF1, red circles) compared to age-and sex-matched controls (CON, grey circles) after controlling for intracranial volume.** Associations are shown for the <u>left</u> hemisphere only. Graphs marked with an *, +, and ° represent significant group-by-age interaction, significant main effect of group, and significant main effect of age respectively. Results remain significant after correcting for multiple comparisons using false discovery rate of *p* = 0.05.

## Notes

### Competing Interest Statement

The authors have declared no competing interest.

### Author Declarations

Ethics committee/IRB of Stanford University School of Medicine gave ethical approval for this work (protocol# 28674).

