## Supplementary material for "Age-related white matter differences in children with neurofibromatosis type 1 compared to controls"

**Table S1.** Mean(M) and standard deviation (SD) of tract-fractional anisotropy (FA) and tract-mean diffusivity (MD) of major white matter pathway in children with neurofibromatosis type 1 (NF1) and age- and sex-matched controls (CON).

|  |  | **Tract-MD** | | **Tract-FA** | |
| --- | --- | --- | --- | --- | --- |
|  | **N** | **NF1**  *M (SD)* | **CON**  *M (SD)* | **NF1**  *M (SD)* | **CON**  *M (SD)* |
| ATR-R | 40 | 0.841 (0.113) | 0.756 (0.055) | 0.389 (0.047) | 0.449 (0.036) |
| ATR-L | 40 | 0.798 (0.127) | 0.727 (0.054) | 0.419 (0.063) | 0.460 (0.039) |
| CST-R | 40 | 0.797 (0.126) | 0.695 (0.060) | 0.582 (0.072) | 0.633 (0.046) |
| CST-L | 40 | 0.789 (0.126) | 0.699 (0.056) | 0.592 (0.072) | 0.646 (0.051) |
| UF-R | 40 | 0.889 (0.110) | 0.819 (0.040) | 0.412 (0.051) | 0.432 (0.041) |
| UF-L | 40 | 0.844 (0.136) | 0.783 (0.045) | 0.435 (0.067) | 0.442 (0.048) |
| Arc-R | 35 | 0.727 (0.097) | 0.693 (0.080) | 0.471 (0.059) | 0.498 (0.060) |
| Arc-L | 40 | 0.818 (0.129) | 0.743 (0.057) | 0.458 (0.070) | 0.494 (0.059) |
| SLF-R | 40 | 0.755 (0.144) | 0.686 (0.065) | 0.459 (0.078) | 0.488 (0.058) |
| SLF-L | 40 | 0.791 (0.128) | 0.718 (0.067) | 0.412 (0.055) | 0.451 (0.063) |
| Cing-R | 35 | 0.832 (0.091) | 0.759 (0.063) | 0.398 (0.063) | 0.431 (0.051) |
| Cing-L | 38 | 0.835 (0.143) | 0.732 (0.063) | 0.454 (0.089) | 0.488 (0.065) |
| IFOF-R | 40 | 0.857 (0.138) | 0.771 (0.068) | 0.467 (0.073) | 0.510 (0.058) |
| IFOF-L | 39 | 0.872 (0.133) | 0.785 (0.063) | 0.458 (0.058) | 0.513 (0.058) |
| ILF-R | 40 | 0.882 (0.140) | 0.810 (0.055) | 0.403 (0.052) | 0.433 (0.050) |
| ILF-L | 40 | 0.899 (0.156) | 0.842 (0.057) | 0.420 (0.059) | 0.436 (0.047) |
| FMinor | 40 | 0.836 (0137) | 0.742 (0.102) | 0.557 (0.059) | 0.633 (0.063) |
| FMajor | 40 | 0.907 (0.183) | 0.800 (0.097) | 0.572 (0.086) | 0.625 (0.053) |

ATR = anterior thalamic radiation, CST = corticospinal tract, Cing = cingulum cingulate, FMajor = forceps major, FMinor = forceps minor, IFOF = inferior fronto-occipital fasciculus, ILF = inferior longitudinal fasciculus, SLF = superior longitudinal fasciculus, UF = uncinate fasciculus, Arc = arcuate fasciculus, L = left / R = right

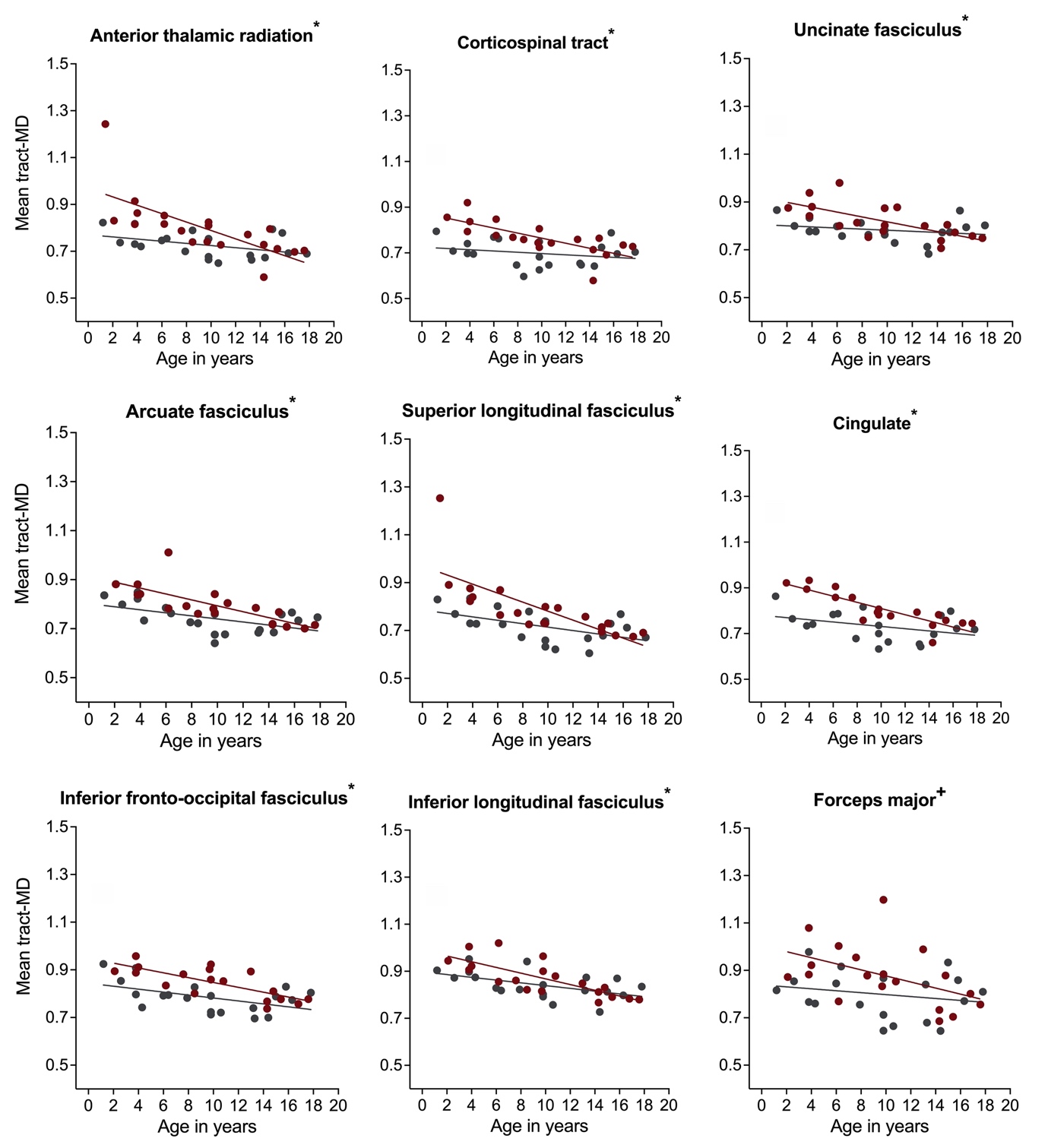

***Figure S1.*** **Associations of mean diffusivity (MD) with age in children with neurofibromatosis type 1 (NF1, red circles) compared to age- and sex-matched controls (CON, grey circles) after controlling for intracranial volume.** Associations are shown for the left hemisphere only. Graphs marked with an *, +, and ° represent significant group-by-age interaction, significant main effect of group, and significant main effect of age respectively. Results remain significant after correcting for multiple comparisons using false discovery rate of *p* = 0.05.

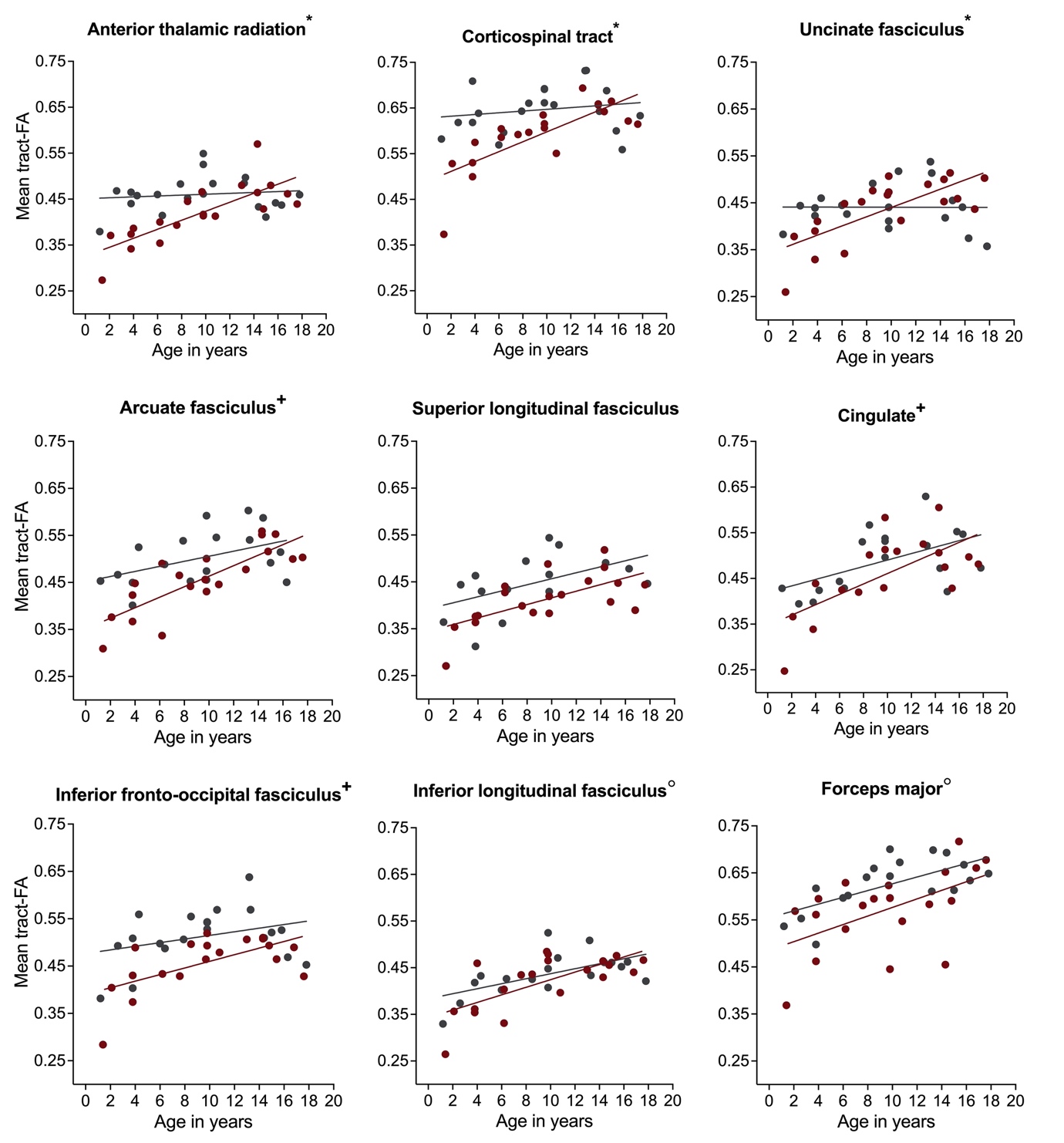

***Figure S2.*** **Associations of fractional anisotropy (FA) with age in children with neurofibromatosis type 1 (NF1, red circles) compared to age- and sex-matched controls (CON, grey circles) after controlling for intracranial volume.** Associations are shown for the left hemisphere only. Graphs marked with an *, +, and ° represent significant group-by-age interaction, significant main effect of group, and significant main effect of age respectively. Results remain significant after correcting for multiple comparisons using false discovery rate of *p* = 0.05.
